# Psychological responses, behavioral changes and public perceptions during the early phase of the COVID-19 outbreak in China: a population based cross-sectional survey

**DOI:** 10.1101/2020.02.18.20024448

**Authors:** Mengcen Qian, Qianhui Wu, Peng Wu, Zhiyuan Hou, Yuxia Liang, Benjamin J. Cowling, Hongjie Yu

**Affiliations:** School of Public Health, Fudan University, Key Laboratory of Public Health Safety, Ministry of Education, Shanghai, China; Key Laboratory of Health Technology Assessment (Fudan University), Ministry of Health, Shanghai, China; World Health Organization (WHO) Collaborating Centre for Infectious Disease Epidemiology and Control, School of Public Health, Li Ka Shing Faculty of Medicine, The University of Hong Kong, Hong Kong Special Administrative Region, China

## Abstract

**Objective:** To investigate psychological and behavioral responses to the threat of SARS-CoV-2 infections and their associations with public perceptions in China

**Design:** Cross sectional population-based telephone survey via random digital dialing between 1 and 10 February, 2020

**Setting:** Wuhan (the epicentre and quarantined city), and Shanghai (a typical major city with close transportation link with Wuhan)

**Participants:** Random sample of 510 residents in Wuhan and 501 residents in Shanghai aged above 18

**Main outcome measures:** Anxiety (measured by the 7-item generalized anxiety disorder [GAD-7] scale), recommended and avoidance behaviors (engaged in all six behaviors such as increasing surface cleaning and reducing going out).

**Results:** The prevalence rates of moderate or severe anxiety (score ≥10 on GAD-7) were 32.7% (n=167) among Wuhan participants and 20.4% (n=102) among Shanghai participants. 78.6% (n=401) of Wuhan participants and 63.9% (n=320) of Shanghai participants had carried out all six precautionary behaviors. For both measures, Wuhan participants were more responsive to the outbreak (p<0.001). Controlling for personal characteristics, logistic regression results suggested that risks of moderate or severe anxiety were positively associated with perceived susceptibility (odds ratio 1.6, 95% confidence interval 1.3-1.8) and severity of the disease (1.6, 1.4-1.9) and confusion about information reliability (1.6, 1.5-1.9). Having confidence in taking measures to protect oneself against the disease was associated with a lower risk (0.6, 0.5-0.7). The strongest predictor of behavioral change was perceived severity (1.2, 1.1-1.4), followed by confusion about information reliability (1.1, 1.0-1.3).

**Conclusions:** Psychological and behavioral responses to COVID-19 have been dramatic during the rising phase of the outbreak. Our results support efforts for timely dissemination of accurate and reliable information to address the high anxiety level.

## 1 Introduction

In December 2019, coronavirus disease 2019 (COVID-19) caused by severe acute respiratory syndrome coronavirus 2 (SARS-CoV-2) emerged in Wuhan city, China.^1^ As of Feb 16, 2020, a total of 70,548 COVID-19 cases with 1,770 deaths had been reported in mainland China.^2^ The outbreak has now spread to twenty-five countries outside of China.^3^ On Jan 31, 2020 (Beijing Time), the World Health Organization (WHO) declared the coronavirus outbreak a public health emergency of international concern (PHEIC). Since Jan 23, 2020, the Chinese government has put Wuhan and several nearby cities under quarantine, hoping to stop the disease from spreading to other parts of the country. However, millions of people had left Wuhan before the lockdown because of the approaching Spring Festival holiday.

Containment measures in the COVID-19 outbreak have focused on identifying, treating, and isolating infected people, tracing and quarantining their close contacts, and promoting precautionary behaviors among the general public. Therefore, the psychological and behavioral responses of the general population play an important role in the control of the outbreak. Previous studies have explored on this topic in various culture settings with SARS,^4,5,6^ pandemic influenza A(H1N1),^7,8,9,10^ and influenza A(H7N9).^11,12,13^ Cultural differences are evident in public responses.^14,15^ Behavioral changes are also associated with government involvement level, perceptions of diseases, and the stage of the outbreak, and these factors vary by diseases and settings. ^4,5,8,16^

The current COVID-19 outbreak provides a unique platform to study behavioral changes for two main reasons. First, government engagement in the control of the outbreak has been unprecedented, for example, locking down Wuhan and surrounding cities, building new hospitals to treat infected patients in Wuhan within two weeks, extending holidays and school closure, deploying thousands of medical staff to heavily affected areas, and running intense public messaging campaigns. Second, the public are faced with rather mixed information, partly because knowledge of the newly emerging disease is evolving with the course of the outbreak. For example, the national technical protocols for COVID-19 released by the National Health Commission have been updated five times within a month. Both features might result in different public responses towards the outbreak. In this study, we aimed to investigate psychological and behavioral responses to the threat of COVID-19 outbreak and to examine public perceptions associated with the response outcomes in mainland China.

## 2 Methods

### 2.1 Cross-sectional telephone survey

A population-based telephone interview was carried out between 1 and 10 February, 2020. The cities of Wuhan and Shanghai were selected to represent diverse exposure intensities to the threat of SARS-CoV-2 infection. Wuhan is the hardest-hit city to date, the city where the virus originated, and the earliest city put under quarantine. By contrast, Shanghai is one of the largest cities in China, and was estimated to have received the largest number of infected travelers from Wuhan.^17,18^ Before our survey, Wuhan confirmed 3,215 cases,^19^ accounting for 27.3% of all cases across China;^20^ Shanghai reported 153 cases, 68 (44.4%) of them from non-local residents.^21^ Figure 1 illustrates the timeline of the outbreak progression compared with our survey dates.

**Figure 1.**
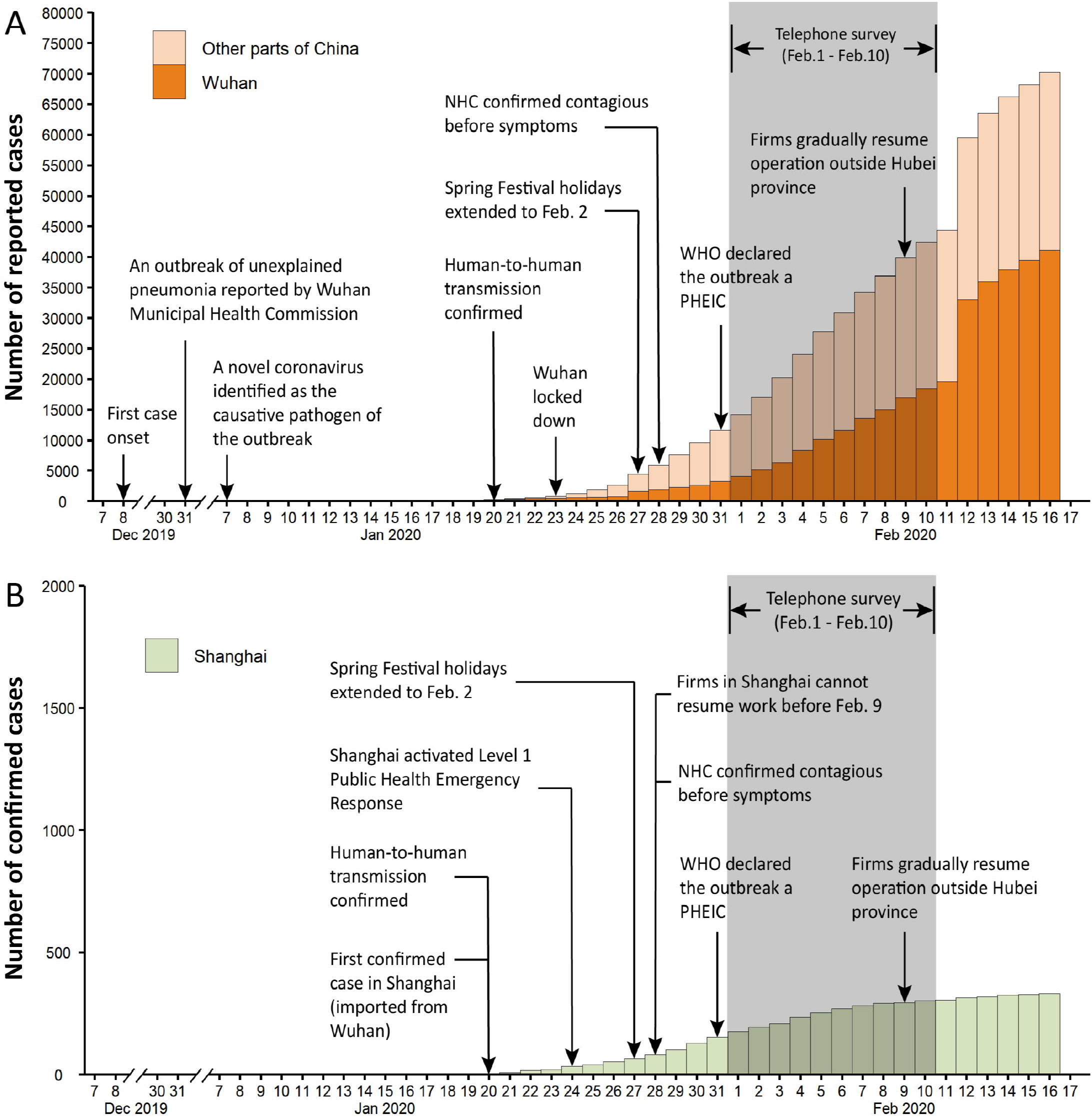
The timeline of the COVID-19 outbreak compared with the survey dates Notes: The timeline of COVID-19 outbreak compared with the survey dates is presented for Wuhan and other parts of China (**Panel A)** and Shanghai (**Panel B**). Note that different scales are used in the two panels. The vertical axis for Panel A is number of reported cases and that for Panel B is number of confirmed cases. Because diagnostic criteria were changed in Hubei Province (Wuhan is the capital city of Hubei Province) on Feb 12, 2020. Since then confirmed cases are based on clinical diagnosis instead of laboratory testing in Hubei province. NHC stands for National Health Commission of China; WHO stands for World Health Organization; PHEIC stands for public health emergency of international concern.

The survey was conducted by well-trained interviewers with 1,011 residents of Wuhan (n=510) and Shanghai (n=501) using a computerized random digital dialing system. The sample size provided us with a sample error of 5%. Proportional quota sampling based on age and sex was used to ensure a demographically representative sample of the general population in each city. Calls were placed three times at different hours on the same day before being classified as invalid. Local residents who were aged 18 years and above and were currently living in the selected cities were eligible to participate. Verbal informed consent was obtained from all participants. Figure S1 presents the flow chart of participant recruitment. Ethical approval was obtained from the institutional review board at School of Public Health, Fudan University (IRB#2020-TYSQ-01-1).

### 2.2 Anxiety

Anxiety was assessed using the 7-item Generalized Anxiety Disorder Scale (GAD-7). The tool is a brief self-reported scale that has demonstrated good reliability and validity in the general population.^22, 23^ Participants were asked to answer how frequently they had been bothered by various symptoms over the past two weeks. The scale accordingly produced a summary GAD scores ranging from 0 to 21. Respondents that scored 10 and above were identified as having moderate or severe level of anxiety.^22^

### 2.3 Behaviors

Participants were asked three questions about recent avoidance behaviors (avoided eating out, avoided taking public transportation, and reduced visits to public places) and another three about recommended behaviors (rescheduling travel plan, increasing surface cleaning, and maintaining better indoor ventilation) in response to the outbreak. All questions were phrased as “during the past week, did you ever … because of the novel coronavirus outbreak”. Possible responses for each question were yes or no.

Three sets of questions were asked to collect information about preventive behaviors in usual days before 31 December 2019 when an unknown pneumonia outbreak related to the later identified SARS-CoV-2 was firstly reported, and then in the past week. The first two sets of questions regarding the frequency of wearing a face mask when going out and the frequency of handwashing immediately when returning home. Response options were never (scored as 1), rare (2), sometimes (3), usually (4), always (5), or did not go out. The top reasons for wearing a face mask in usual days and in the past week were collected. Respondents who did not always wear a face mask in the past week were also required to provide a most possible reason. The third set of question concerning handwashing duration with five responses categories from short to long (1 score to 5 scores), for less than 10 seconds, 10-19 seconds, 20-39 seconds, 40-59 seconds, or 60 seconds and above, respectively. Respondents whose answers for behaviors in the past week scored higher than those in usual days were categorized as increased frequency or duration of the aforementioned behaviors.

To capture possible over-reaction of the public, a supplementary question was asked “In the past week, have you ever purchased or tried to purchase goggles for the purpose of protection against infection with SARS-CoV-2?” with two possible responses (yes or no).

### 2.4 Perceptions

Five items were used to assess whether participants believed that certain measures would reduce their risk of catching COVID-19, with possible response options being ineffective (1), even (2), or effective (3). The first three items (frequent handwashing, wearing a face mask, and avoiding going out) were consistent with the advertising campaign run by the government. The last two items (washing mouth with salty water, and taking vitamin C or a product of the traditional Chinese medicine, isatis root) were considered as misinformation by medical experts. Participants answered effective in all first three items and ineffective in both of the last two items were categorized as having correct perceived efficacy of behaviors.

Four items assessed perceived threats of the novel coronavirus. Participants were asked “how likely do you think you will contract novel coronavirus infection over the next month” with five responses from very unlikely (1) to very likely (5), and “how serious do you think the novel coronavirus infection would be if you contracted it” with five options from very mild (1) to very serious (5). Participants were also required to report relative transmissibility and severity compared with SARS in 2003 with five response categories being much lower (1), lower (2), similar (3), higher (4), or much higher (5).

Three items assessed how well informed were the public. Participants were asked “whether you have received and read the information brochure about the novel coronavirus from the government and medical experts” with two response options (yes or no). They were then required to provide response to the statement “information that I received about the novel coronavirus outbreak is sufficient” with five options from strongly disagree (1) to strongly agree (5). To assess the impact of mixed information during the outbreak, participants were asked to report the frequency that they felt confused or bothered about the reliability of the information that they received. Response options ranged from never (1) to always (5).

The confidence of the public was assessed by asking if they agreed with the statement “I believe that I can take measures to protect myself against the novel coronavirus” with five response options from strongly disagree (1) to strongly agree (5).

### 2.5 Personal characteristics

Personal characteristics were collected in the following dimensions: sex, age, working status, perceived household income level, whether experienced symptoms (cough and fever) during the past two weeks, whether has friends or relatives with symptoms in the past two weeks, and whether there had been confirmed or suspected cases of novel coronavirus infections in their neighborhood.

### 2.6 Analyses

Analyses were carried out in Stata version 14 for Windows. Descriptive analysis of the aforementioned responses was performed to report the counts and frequencies, stratified by cities. Differences across cities were examined using t tests for dummy variables and Chisquare or fisher’s exact tests (when applicable) for variables measured by a Likert scale. The primary outcomes were moderate or severe anxiety (GAD-7 score ≥10), and whether respondents engaged in all recommended and avoidance behaviors. Multivariate logistic regression models were first used to estimate associations between personal characteristics and primary outcomes. These models adjusting for personal characteristics were then used to investigate associations between perception variables and primary outcomes. Perception variables were included in the form of continuous scores.

### 2.7 Patient and public involvement

Public were involved in this study as participants of the telephone survey.

## 3 Results

### 3.1 Sample characteristics

Overall, 18,576 randomly generated digital numbers were selected and called. Of these, 7,341 calls answered; of these, 5,650 declined the participation invitation, 505 were ineligible, 185 dropped before completion, leaving 1,011 respondents successfully completed the interview. The response rate was 13.8% (1,011/7,341) (Figure S1). Participants in Wuhan (34.5%) reported a much higher rate of living in neighborhood with confirmed or suspected novel coronavirus cases than those in Shanghai (6.4%) (Table 1).

**Table 1.** Personal characteristics and associations with psychological and behavioral responses

|  | Wuhan | Shanghai | Moderate or severe anxiety | Reported all recommended and avoidance behaviors |
| --- | --- | --- | --- | --- |
|  | N (%) | N (%) | OR (95% CI) | OR (95% CI) |
| Male | 255 (50.0) | 255 (50.9) | 1.0 (0.7 - 1.3) | 0.8 (0.6 - 1.1) |
| Age, years |  |  |  |  |
| 18-24 | 89 (17.5) | 75 (15.0) | Reference | Reference |
| 25-39 | 161 (31.6) | 176 (35.1) | 0.7 (0.4 - 1.2) | 0.7 (0.4 - 1.1) |
| 40-59 | 197 (38.6) | 165 (32.9) | 1.2 (0.7 - 2.2) | 0.8 (0.4 - 1.4) |
| 60+ | 63 (12.4) | 85 (17.0) | 1.2 (0.6 - 2.4) | 0.7 (0.4 - 1.5) |
| Educational attainment |  |  |  |  |
| Junior school and below | 27 (5.3) | 38 (7.6) | Reference | Reference |
| High school equivalent | 110 (21.6) | 72 (14.4) | 2.2 (1.0 - 5.0) | 1.1 (0.6 - 2.0) |
| College equivalent | 355 (69.6) | 337 (67.3) | 2.1 (0.9 - 5.1) | 1.8 (0.9 - 3.8) |
| Graduate and above | 18 (3.5) | 54 (10.8) | 2.5 (0.9 - 6.5) | 1.2 (0.5 - 2.6) |
| Working status |  |  |  |  |
| Unemployed/retired | 148 (29.0) | 142 (28.3) | Reference | Reference |
| Employed | 362 (71.0) | 359 (71.7) | 1.0 (0.7 - 1.5) | 1.0 (0.6 - 1.5) |
| Married | 383 (75.1) | 344 (68.7) | 0.7 (0.4 - 1.1) | 1.1 (0.7 - 1.7) |
| Income |  |  |  |  |
| Below median | 116 (22.7) | 97 (19.4) | Reference | Reference |
| Median | 329 (64.5) | 306 (61.1) | 0.6** (0.4 - 0.9) | 1.2 (0.9 - 1.8) |
| Above median | 65 (12.7) | 98 (19.6) | 0.6 (0.4 - 1.1) | 0.8 (0.5 - 1.3) |
| Experienced symptoms | 63 (12.4) | 51 (10.2) | 1.3 (0.9 - 2.1) | 0.6* (0.4 - 1.0) |
| Friends with symptoms |  |  |  |  |
| No | 437 (85.7) | 464 (92.6) | Reference | Reference |
| Yes | 63 (12.4) | 36 (7.2) | 0.7 (0.4 - 1.2) | 0.7 (0.4 - 1.1) |
| Don't know | 10 (2.0) | 1 (0.2) | 0.3 (0.0 - 1.6) | 1.1 (0.3 - 4.1) |
| Suspected cases in neighborhood |  |  |  |  |
| No | 261 (51.2) | 413 (82.4) | Reference | Reference |
| Yes | 176 (34.5) | 32 (6.4) | 1.9** (1.3 - 2.8) | 0.6* (0.4 - 0.9) |
| Don't know | 73 (14.3) | 56 (11.2) | 1.5 (1.0 - 2.4) | 0.9 (0.6 - 1.5) |
| City |  |  |  |  |
| Wuhan | -- | -- | Reference | Reference |
| Shanghai | -- | -- | 0.6** (0.5 - 0.8) | 0.4** (0.3 - 0.6) |

Notes: The first two columns report counts and frequencies of each variable value. The last two column present multivariate regression results. \* for significance at the 5% level; \*\* for significance at the 1% level.

### 3.2 Anxiety, behavioral outcomes and perceptions

A total of 167 (32.7%) respondents in Wuhan and 102 (20.4%) respondents in Shanghai were identified to have moderate or severe anxiety (Panel A Table 2). The prevalence was significantly higher in Wuhan (p<0.001). The mean GAD-7 scores were 7.1 and 5.1 in Wuhan and Shanghai, respectively (p<0.001).

**Table 2.** Anxiety, behavioral responses and perceptions during the novel coronavirus outbreak in Wuhan and Shanghai

|  | Wuhan<br>(N=510)<br>N (%) | Shanghai<br>(N=501)<br>N (%) | P-value |
| --- | --- | --- | --- |
| **Panel A: Anxiety levels** |  |  |  |
| Moderate or severe anxiety<br>(GAD-7 score $\geq 10$) | 167 (32.7) | 102 (20.4) | <0.001 |
| **Panel B: Behavioral outcomes** |  |  |  |
| Engaged in all recommended and avoidance behaviors | 401 (78.6) | 320 (63.9) | <0.001 |
| Never went out last week | 124 (24.3) | 69 (13.8) | <0.001 |
| More often wore a face mask when going out<sup>1</sup> | 355 (92.0) | 373 (86.3) | 0.853 |
| More often washed hands immediately<br>when returning home<sup>1</sup> | 164 (42.5) | 157 (36.3) | 0.073 |
| Longer hand washing duration | 260 (51.0) | 200 (39.9) | <0.001 |
| Purchased goggles for prevention | 143 (28.0) | 105 (21.0) | 0.009 |
| **Panel C: Perception variables** |  |  |  |
| Correct perceived efficacy of behaviors | 65 (12.7) | 97 (19.4) | 0.004 |
| Self-perceived risk<sup>2</sup> |  |  | 0.033 |
| Very unlikely | 85 (18.2) | 93 (19.7) |  |
| Unlikely | 190 (40.7) | 230 (48.6) |  |
| Even | 105 (22.5) | 91 (19.2) |  |
| Likely | 72 (15.4) | 49 (10.4) |  |
| Very likely | 15 (3.2) | 10 (2.1) |  |
| Self-perceived severity if contracted<sup>3</sup> |  |  | <0.001 |
| Very mild | 158 (39.7) | 104 (25.8) |  |
| Mild | 142 (35.7) | 166 (41.2) |  |
| Moderate | 50 (12.6) | 53 (13.2) |  |
| Serious | 32 (8.0) | 60 (14.9) |  |
| Very serious | 16 (4.0) | 20 (5.0) |  |
| Transmissibility compared with SARS |  |  | <0.001 |
| Much lower | 13 (2.5) | 1 (0.2) |  |
| Lower | 17 (3.3) | 8 (1.6) |  |
| Even | 59 (11.6) | 39 (7.8) |  |
| Higher | 170 (33.3) | 172 (34.3) |  |
| Much higher | 251 (49.2) | 281 (56.1) |  |
| Harm to body compared with SARS |  |  | 0.370 |
| Much lower | 13 (2.5) | 12 (2.4) |  |
| Lower | 66 (12.9) | 75 (15.0) |  |
| Even | 131 (25.7) | 136 (27.1) |  |
| Higher | 158 (31.0) | 127 (25.3) |  |
| Much higher | 142 (27.8) | 151 (30.1) |  |
| Received and read information brochures | 490 (96.1) | 446 (89.0) | <0.001 |
| Sufficient information |  |  | 0.937 |
| Strongly disagree | 4 (0.8) | 7 (1.4) |  |
| Disagree | 23 (4.5) | 23 (4.6) |  |
| Even | 65 (12.7) | 65 (13.0) |  |
| Agree | 221 (43.3) | 221 (44.1) |  |
| Strongly agree | 197 (38.6) | 197 (39.3) |  |
| Confused about information reliability |  |  | 0.003 |
| Never | 158 (31.0) | 215(42.9) |  |
| Rare | 150 (29.4) | 121(24.2) |  |
| Sometimes | 124 (24.3) | 102(20.4) |  |
| Usually | 46 (9.0) | 41(8.2) |  |
| Always | 32 (6.3) | 22(4.4) |  |
| Confidence on taking measures to protect myself |  |  | 0.302 |
| Strongly disagree | 2 (0.4) | 3 (0.6) |  |
| Disagree | 13 (2.5) | 14 (2.8) |  |
| Even | 47 (9.2) | 49 (9.8) |  |
| Agree | 253 (49.6) | 276 (55.1) |  |
| Strongly agree | 195 (38.2) | 159 (31.7) |  |

<sup>1</sup> The total number of respondents was 386 in the Wuhan sample and 432 in the Shanghai sample because 124 and 69 participants reported “never went out last week” respectively in the above samples.
<sup>2</sup> The total number of respondents was 457 in the Wuhan sample and 473 in the Shanghai sample because 43 Wuhan participants and 28 Shanghai participants refused to answer.
<sup>3</sup> The total number of respondents was 398 in the Wuhan sample and 403 in the Shanghai sample because 112 Wuhan participants and 98 Shanghai participants refused to answer.

In total 401 (78.6%) Wuhan participants and 320 (63.9%) Shanghai participants engaged in all six recommended and avoidance behaviors due to the novel coronavirus outbreak (Panel B Table 2). A high compliance rate of above 90% was observed in all other behaviors in both samples, except for increased surface cleaning, which was driving the significant observed differences in behavioral responses between the two cities (Table S1). Frequencies of washing hands immediately when returning home, wearing a face mask when going out, and duration of handwashing significantly increased in both cities during the outbreak (Panel B Table 2; Figure 2). We found no statistically significant differences in changes of the first two aforementioned behaviors. By contrast, more respondents reported longer duration of handwashing in Wuhan (p<0.001). Moreover, a significantly higher proportion of Wuhan respondents reported purchasing or trying to purchase goggles for prevention purpose (p=0.009).

**Figure 2.**
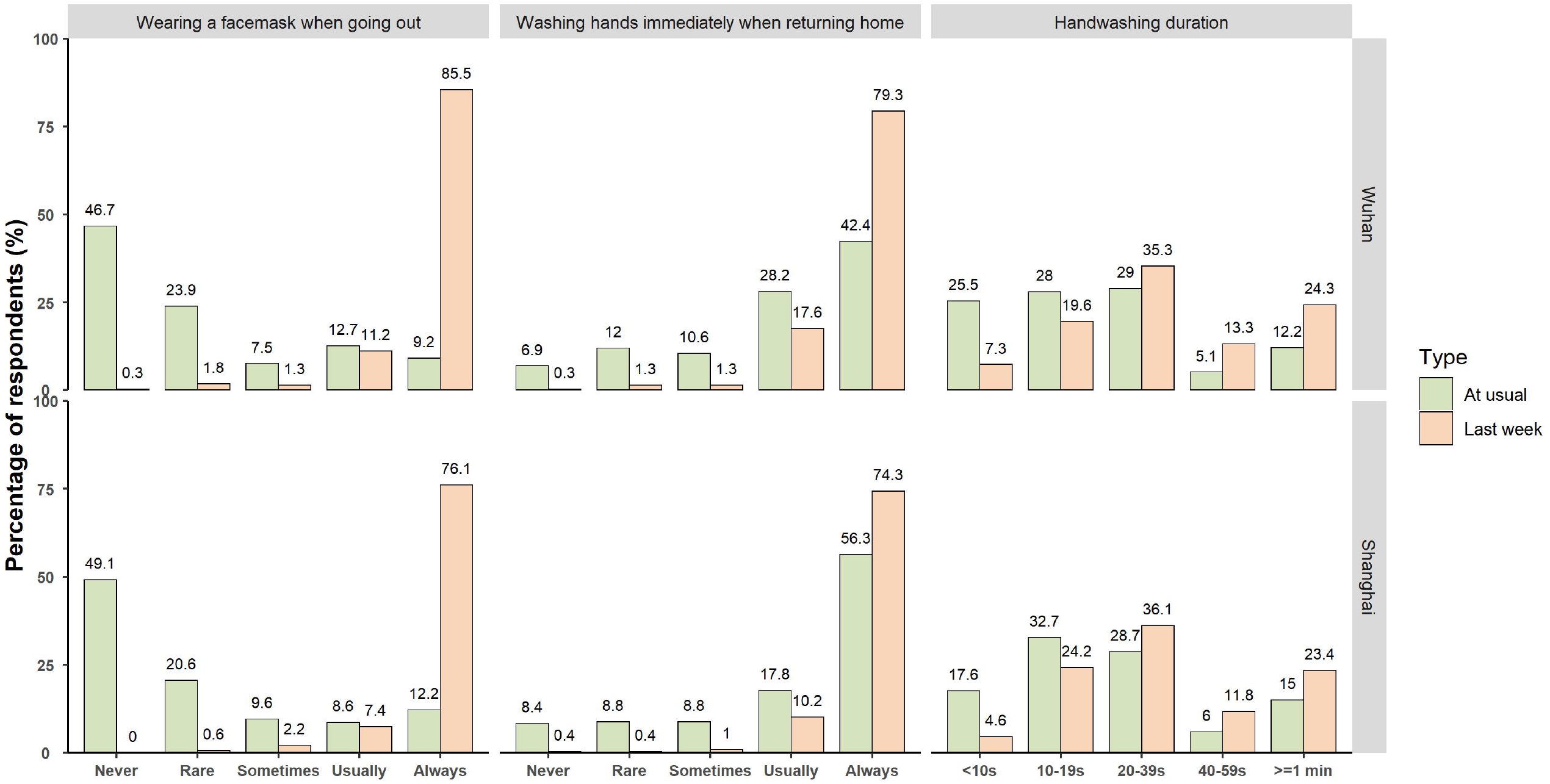
Handwashing and face mask wearing behaviors in usual days and during the past week in Wuhan and Shanghai.

Overall, 12.7% Wuhan participants responded with correct perceived efficacy of behaviors. The proportion was significantly higher (p=0.004) in the Shanghai sample (19.4%) (Panel C Table 2). Generally, Wuhan respondents reported significantly higher self-perceived risk (p=0.033) of contracting novel coronavirus than their counterparts in Shanghai; however, their perceived severity and relative transmissibility to SARS were significantly lower (p<0.001). No differences were found between the two cities in terms of perceived harm to body compared with SARS. Moreover, respondents aged 40 and above, who presumably have clearer memory of SARS, had higher (p<0.001) perceived relative severity than those aged 18-39 in Shanghai; similar findings were not found in Wuhan (Table S2).

Around 96.1% Wuhan respondents had received and read the information brochures of the novel coronavirus from the government or medical experts. The proportion was significantly lower (p<0.001) but still very high in Shanghai (89.0%). The majority of the respondents in both cities thought that the information that they received was sufficient, without discernable differences. By contrast, the Wuhan respondents were significantly more often confused about the reliability of the information that they received (p=0.003); only 31% respondents in Wuhan never felt bothered about this issue, while the corresponding figure in Shanghai was 42.9%. The majority of the participants agreed or strongly agreed with having confidence on taking measures to protect themselves. We detected no statistically significant differences across cities in this domain.

### 3.3 Personal variables associated with psychological and behavioral responses

The largest positive and significant effects on moderate or severe anxiety were from participants with confirmed or suspected cases in their living neighborhood (odds ratio 1.9, 95% confidence interval 1.3-2.8) (Table 1). Compared to low income individuals, those with median income were less likely to suffer moderate or severe anxiety (odds ratio 0.6, 0.4-0.9). Furthermore, Shanghai participants were significantly less likely to report moderate or severe anxiety (odds ratio 0.6, 0.5-0.8) or carry out all recommended and avoidance behaviors (odds ratio 0.4, 0.3-0.6) than Wuhan participants. We found no evidence that sex, age, educational attainment, working status and marital status were closely associated with psychological and behavioral responses during the novel coronavirus outbreak.

### 3.4 Perception variables associated with psychological and behavioral responses

We found that higher perceived risk and severity of contracting the novel coronavirus, higher perceived relative transmissibility and harm to body to SARS, and more confusion about information reliability were all significantly and positively associated with reported moderate or severe anxiety during the outbreak (Table 3). In contrast, stronger self-confidence was significantly associated with lower risks of moderate or severe anxiety.

**Table 3.**
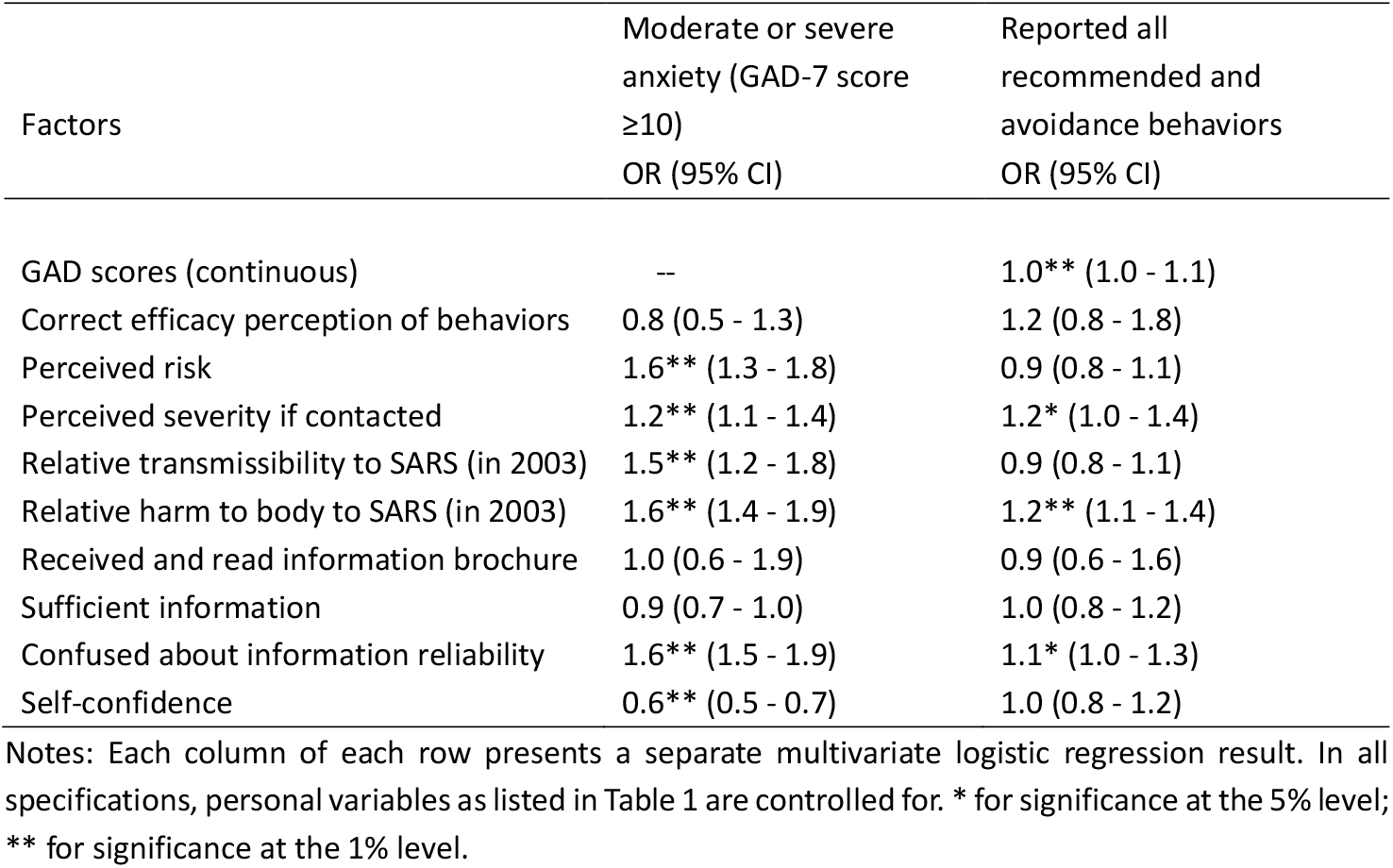
Perception factors associated with anxiety and behavioral responses

For behavioral responses, increases in anxiety levels, perceived risks and harms to body relative to SARS, and confusion about information reliability were significantly associated with higher probability of carrying out all recommended and avoidance behaviors during the outbreak (Table 3). For increases in frequencies of immediate handwashing behavior when returning home, the factor that exerted the largest impact was having received and read information brochures from the government, followed by having correct perceived efficacy of behaviors (Table S3). Higher perceived harm to body compared with SARS was the only factor with significant explanatory power for goggles purchase behavior. We found no evidence that perceptions and anxiety levels were associated with changes in frequencies of wearing face masks during the outbreak (Table S3).

## 4 Discussion

We conducted a population-based telephone survey via random digital dialing to investigate anxiety levels, changes in precautionary behaviors and public perceptions among Wuhan and Shanghai residents during the early phase of 2019 novel coronavirus. Around of 32.7% Wuhan and 20.4% Shanghai participants reported moderate or severe anxiety; 78.6% Wuhan and 63.9% Shanghai respondents carried out all six precautionary behaviors due to the outbreak. Wuhan residents were significantly more responsive than their Shanghai counterparts in the two aforementioned outcomes. Our results suggested that perceived risk and severity of the disease as well as information reliability were important factors associated with the psychological and behavioral responses.

Our study provides new evidence on psychological and behavioral response to disease outbreaks by comparing public response in two cities under various exposures to the early phase of the COVID-19 outbreak. The results are an important addition to previous literature in several dimensions. First, the findings suggest that anxiety levels and precautionary behaviors have changed rapidly and dramatically in both studied cities during the early stage of the outbreak, regardless of their quarantine status. The prevalence of moderate or severe anxiety has been 4-5 times of its normal level in urban China; ^24,25^ the percentage of always wearing a face mask surged from 9.2-12.2% in usual days to 76.1-85.5 during the past week (Figure 2); preventing illness and virus has been the most evident reason for such a drastic change (Figure S2); and the main reason for those not always wearing a facemask after the outbreak was the face-mask shortage. These results contradict to findings in UK during the influenza A(H1N1)pdm09 pandemic,^7^ but are consistent with and much sizeable than those in Hong Kong during SARS and influenza A(H1N1). ^4,8,10^

Second, our results suggest that more responsive behavioral changes observed in this study were associated with the strong involvement from the Chinese government during the control and prevention of coronavirus than voluntary avoidance behaviors reported previously. ^5,9,26,27,^ Measures such as temporary closing down public transportation in Wuhan and imposing punishment for passengers not wearing a face mask on metro in Shanghai have fueled the high compliance rates with government recommendations (Table S1) and perceived efficacy of these behaviors (Table S4). These polices are also responsible for the limited observed heterogeneity in uptake and changes of precautionary behaviors. For example, we find that no perception variables exerted a statistically meaningful effect on increased face mask wearing frequency (Table S3).

Third, we provided evidence for unwarranted precautionary behavior in coping with a novel disease, which was less documented in the literature.^5^ Around 28% Wuhan and 21% Shanghai respondents reported having purchased or having tried to purchase goggles for infection prevention. According to the National Health Commission experts, goggles are unnecessary protective equipment for people other than medical staff on the front line of coronavirus outbreak. However, goggles were sold out nationwide shortly after a doctor wrote on social media and suspected that lack of eye protection in early investigation of the outbreak might have led to his infection. Our results showed that the irrational behavior was mainly driven by perceived higher severity than SARS and the over-reaction was more prevalent in Wuhan (Table S3).

Fourth, perceived susceptibility and severity of the coronavirus were higher compared with studies regarding other diseases. After translating our results into comparable scales, we found that the perceived chance of infection (12.5-18.6%) was higher than that during H7N9 in urban China (1.0-2.6%),^11^ and SARS in Hong Kong (3.9-14.3%);^4^ and the relative severity to SARS was four times higher than that of H7N9 in urban China (8.9-11.4%).^11^ Though higher perceived risk and harm of coronavirus positively associated with favorable responses in behaviors, it also led to significantly higher anxiety levels among the general public.

Our findings also yield several important public implications. First, providing the public with reliable and accurate information is crucial for addressing the psychological effects of contagious disease outbreaks. Public confusion about the reliability of information that they received might have led to significantly sustained anxiety. About 57.1-69.0% individuals were ever bothered by this issue, and the prevalence of this issue was significantly more common among people subject to quarantine in Wuhan, suggesting stronger desire for facts.^28^ Second, consequences of misinformation could be long-lasting and should not be underestimated in health crisis management.^7,29^ Although efficacy of behaviors supported in advertising campaign was well received, rumors that have been proved wrong earlier during the SARS outbreak, for example, washing mouth with salty water, or having vitamin C or radix isatidis, still found their audience. Only 24.6-37.9% respondents in our study correctly considered these measures as ineffective (Table S4); the correct rate was significantly lower in Wuhan. Finally, awareness of hand hygiene in China has been low, which calls for more handwashing education in the community. We found that the proportion of handwashing duration over 40 seconds as recommended by WHO only slightly increased from 17.3% to 37.6% during novel coronavirus outbreak.^30^ Fortunately, having received and read information brochures was positively associated with longer handwashing duration (Table S3), suggesting education programs will be effective for addressing this issue.

Our study has a number of limitations. First, in order to investigate public response rates in the immediate aftermath of the major public health crisis, we shortened our survey questionnaire and chose to use random digital dialing with quota sampling to obtain demographically representative samples for both selected cities. We deliberately selected out times of handwashing as a key behavior measure, although it is widely discussed in the literature. Because people have largely reduced going out during the extended national holidays while embracing remote work, resulting in less times of handwashing than in usual days, which makes this measure less valid. Second, we asked respondents to recall some of their behaviors in usual days before December 2019. Their answers might suffer recall bias.

## 5 Conclusions

In conclusion, it is seen that psychological and behavioral responses to COVID-19 had been dramatic during the rising phase of the outbreak. Prevalence of moderate or severe anxiety were 4-5 times of its normal level in urban China. The majority engaged in all six recommended and avoidance behaviors. Wuhan, the epicentre and quarantined city, was significantly more responsive in the aforementioned domains than Shanghai. Tremendous government efforts in control and prevention of the disease were associated with sizable changes in face mask wearing. However, confusion about information reliability significantly fueled the public anxiety levels, and public awareness of hand hygiene was less optimal. Our results support efforts for timely dissemination of accurate and reliable information and to focus more on handwashing education.

## Data Availability

Full top line results for the survey are available from H.Y. at.

## Competing interest statement

H.Y. has received research funding from Sanofi Pasteur, GlaxoSmithKline, Yichang HEC Changjiang Pharmaceutical Company and Shanghai Roche Pharmaceutical Company. None of that research funding is related to COVID-19. BJC has received honoraria from Roche and Sanofi. All other authors report no competing interests. All authors have completed the Unified Competing Interest form.

## Contributors

H.Y. conceptualized the study design. M.Q., Z.H, and Y.L. developed the survey questionnaire and collected data. M.Q. and Q.W. analyzed data. M.Q. interpreted results and wrote the manuscript. P.W., B.J.C., and H.Y. edited the manuscript.

## Transparency declaration

A statement that the lead author, H.Y., affirms that the manuscript is an honest, accurate, and transparent account of the study being reported; that no important aspects of the study have been omitted.

## Ethical approval

Ethical approval was obtained from the institutional review board at Fudan University School of Public Health (IRB#2020-TYSQ-01-1).

## Role of the funding source

H.Y. acknowledges financial support from the National Science Fund for Distinguished Young Scholars (No. 81525023), Key Emergency Project of Shanghai Science and Technology Committee (No. 20411950100), National Science and Technology Major Project of China (No. 2018ZX10201001-010, No. 2017ZX10103009-005, No. 2018ZX10713001-007). M.Q. acknowledges financial support from the National Natural Science Foundation of China (No. 71704027) and Shanghai Municipal Education Commission and Shanghai Education Development Found for Chenguang Program (No. 17CG03). The funders played no part in the study design; in the collection, analysis, and interpretation of data; in the writing of the report; and in the decision to submit the article for publication. The views expressed in this publication are those of the authors and not necessarily those of their funders or employers.

## Acknowledgements

We thank Mark Jit from London School of Hygiene and Tropical Medicine for his valuable comments. We are thankful to Xinyu Wang, Shengyi Pan, Zihan Xu, Longfei Feng, Kaiyue Ren, Yifan Cheng, Abudukelimu Nazhakaiti, Zhiqiang Qu, Geshu Zhang, Ji Geer Guliyeerke, Qian Lv, Biao Wang, LinQing Zhou, and Cong Ma from School of Public Health, Fudan University for providing assistance with data collection.

## Data sharing statement

Full top line results for the survey are available from H.Y. at.

## References

1. Zhu N, Zhang D, Wang W, Li X, Yang B, Song J, et al. A Novel Coronavirus from Patients with Pneumonia in China, 2019. New England Journal of Medicine 2020; DOI: 10.1056/NEJMoa2001017.

2. National Health Commission. Latest situation of new coronavirus pneumonia as of February 16 2020 (in Chinese); (http://www.nhc.gov.cn/xcs/yqfkdt/202002/18546da875d74445bb537ab014e7a1c6.shtml.).

3. World Health Organization. Coronavirus disease 2019 (COVID-19) Situation Report - 26. 2020; (https://www.who.int/docs/default-source/coronaviruse/situation-reports/20200215-sitrep-26-covid-19.pdf?sfvrsn=a4cc6787_2.).

4. Lau JT, Yang X, Tsui H, Kim JH, et al. Monitoring community responses to the SARS epidemic in Hong Kong: form day 10 to day 62. Journal of Epidemiology and Community Health 2003; 57(11): 864–870.

5. Brug J, Ar AR, Oenema A, de Zwart O, Richardus JH, Bishop GD. SARS risk perception, knowledge, precautions, and information sources, the Netherlands. Emerging Infectious Diseases 2004; 10(8): 1486–1489.

6. Wu P, Fang Y, Guan Z, Fan B, Kong J, Yao Z, et al. The psychological impact of the SARS epidemic on hospital employees in China: exposure, risk perception, and altruistic acceptance of risk. Can J Psychiatry 2009; 54(5): 302–311.

7. Rubin GJ, AmlÔt R, Page L, Wessley S. Public perceptions, anxiety, and behaviour change in relation to the swine flu outbreak: cross sectional telephone survey. BMJ 2009; 339: b2651.

8. Yeung NCY, Lau JTF, Choi KC, Griffiths S. Population responses during the pandemic phase of the influenza A(H1N1)pdm09 Epidemic, Hong Kong, China. Emerging Infectious Diseases 2017; 23(5): 813–815.

9. Bayham J, Kuminoff NV, Gunn Q, Fenichel EP. Measured voluntary avoidance behaviour during the 2009 A/H1N1 epidemic. Proceedings of the Royal Society B: Biological Sciences 2015; 282(1818): 20150814.

10. Lau JT, Griffiths S, Choi KC, Tsui HY. Avoidance behaviors and negative psychological responses in the general population in the initial stage of the H1N1 pandemic in Hong Kong. BMC Infectious Diseases 2020; 10: 139.

11. Wang L, Cowing B, Wu P, Yu J, Li F, Zeng L, et al. Human exposure to live poultry and psychological and behavioral responses to influenza A(H7N9), China. Emerging Infectious Diseases 2014; 20(8): 1296–1305.

12. Wu P, Wang L, Cowling BJ, Yu J, Fang VJ, Li F, et al. Live poultry exposure and public response to influenza A(H7N9) in urban and rural China during two epidemic waves in 2013-2014. PLoS ONE 2015; 10(9): e0137831.

13. Wu P, Fang V, Liao Q, Ng DMW, Wu J, Leung GM, et al. Responses to threat of influenza A(H7N9) and support for live poultry markets, Hong Kong, 2003. Emerging Infectious Diseases 2014; 20(5): 882–886.

14. Cheng C, Tang CS. The psychology behind the masks: psychological responses to the severe acute respiratory syndrome outbreak in different regions. Asian Journal of Social Psychology 2004; 7: 3–7.

15. Vartti A-M, Oenema A, Schreck M, Uutela A, de Zwart O, Brug J, et al. SARS knowledge, perceptions, and behaviors: a comparison between Finns and the Dutch during the SARS outbreak in 2003. Int J Behav Med 2009; 16:41–18.

16. de Zwart O, Veldhuijzen IK, Elam G, Aro AR, Abraham T, Bishop GD, et al. Perceived threats, risk perception, and efficacy beliefs related to SARS and other (emerging) infectious diseases: results of an international survey. International Journal of Behavioral Medicine 2009; 16: 30–40.

17. Wu J, Leung K, Leung GM. Nowcasting and forecasting the potential domestic and international spread of the 2019-nCoV outbreak originating in Wuhan, China: a modelling study. Lancet 2020; https://doi.org/10.1016/S0140-6736(20)30260-9.

18. Read JM, Bridgen JRE, Cummings DAT, Ho A, Jewell CP. Novel coronavirus 2019-nCoV: early estimation of epidemiological parameters and epidemic predictions. Preprint at https://www.medrxiv.org/content/10.1101/2020.01.23.20018549v2 (2020).

19. Health Commission of Hubei Province. Update on the outbreak of novel coronavirus-infected pneumonia as of 31 January 2020. http://wjw.hubei.gov.cn/fbjd/dtyw/202002/t20200201_2017100.shtml (accessed on February 15, 2020) (in Chinese).

20. National Health Commission of the People’s Republic of China. Update on the outbreak of novel coronavirus-related pneumonia as of 31 January 2020. http://www.nhc.gov.cn/xcs/yqtb/202002/84faf71e096446fdb1ae44939ba5c528.shtml (accessed on February 15, 2020) (in Chinese).

21. Shanghai Municipal Health Commission, Shanghai Municipal Administration of Traditional Chinese Medicine. 18 new confirmed cases of novel coronavirus-infected pneumonia in Shanghai. http://wsjkw.sh.gov.cn/yqtb/20200201/c109479681e64952824c04eae3145e89.html (accessed on February 15, 2020) (in Chinese).

22. Spitzer RL, Kroneke K, Williams JBW, Löwe B. A brief measure for assessing generalized anxiety disorder. Archives of Internal Medicine 2006; 166: 1092–1097.

23. Löwe B, Decker O, Müller S, Brähler E, Schellberg D, Wolfgang H, et al. Validation and standardization of the Generalized Anxiety Disorder Screener (GAD-7) in the general population. Medical Care 2008; 46(3): 266–274.

24. Guo X, Meng Z, Huang G, Fan J, Zhou W, Ling W, et al. Meta-analysis of the prevalence of anxiety disorders in mainland China from 2000 to 2015. Scientific Reports 2016; 6: 28033.

25. Yu W, Singh SS, Calhoun S, Zhang H, Zhao X, Yang F. Generalized anxiety disorder in urban China: prevalence, awareness, and disease burden. Journal of Affective Disorder 2018; 234: 89–96.

26. Fenichel EP, Kuminoff NV, Chowell G. Skip the trip: air travelers’ behavioral responses to pandemic influenza. PLoS ONE 2013; 8(3): e58249.

27. Sadique MZ, Edmunds WJ, Smith RD, Merrding WJ, de Zwart O, Brug J. Precautionary behavior in response to perceived threat of pandemic influenza. Emerging Infectious Disease 2007; 13(9): 1307–1313.

28. Rubin GJ, Wessely S. Coronavirus: the psychological effects of quarantining a city. BMJ 2020; https://blogs.bmj.com/bmj/2020/01/24/coronavirus-the-psychological-effects-of-quarantining-a-city/

29. Qian M, Chou S, Lai EK. Confirmatory bias in health decisions: evidence form the MMR-autism controversy. Journal of Health Economics 2020; https://doi.org/10.1016/j.jhealeco.2019.102284

30. World Health Organization. Hand hygiene: why, how & when? https://www.who.int/gpsc/5may/Hand_Hygiene_Why_How_and_When_Brochure.pdf (accessed on February 15, 2020).

